# Imaging protocol suggested by large language model depends on language: preliminary experiments using GPT-4

**DOI:** 10.1101/2024.07.31.24311123

**Authors:** Kazufumi Suzuki, Kayoko Abe, Shuji Sakai

## Abstract

**Purpose:** This study aimed to evaluate the potential of GPT-4, a large language model, in assisting radiologists to determine brain magnetic resonance imaging (MRI) protocols.

**Methods:** We used brain MRI protocols from a specific hospital, covering 20 diseases or examination purposes, excluding brain tumor protocols. GPT-4 was given system prompts to add one MRI sequence for the basic brain MRI protocol and disease names were input as user prompts. The model’s suggestions were evaluated by two radiologists with over 20 years of relevant experience. Suggestions were scored based on their alignment with the hospital’s protocol as follows: 0 for inappropriate, 1 for acceptable but nonmatching, and 2 for matching the protocol. The experiment was conducted in both Japanese and English to compare GPT-4’s performance in different languages.

**Results:** GPT-4 scored 27/40 points in English and 28/40 points in Japanese. GPT-4 gave inappropriate suggestions for Moyamoya disease and neuromyelitis optica in both languages and cerebral infarction in Japanese. For the other protocols, the suggested sequences were either appropriate or better. The suggestions in English differed from those in Japanese for seven protocols.

**Conclusion:** GPT-4 can suggest appropriate MRI sequences for each disease in addition to the standard brain MRI protocol. GPT-4’s output is language-dependent and suggests brain MRI protocols tailored to specific regions and domains.

## Introduction

The imaging methods of MRI vary depending on the disease under investigation, and their determination is entrusted to radiologists. However, this task is often extensive and complex. In this study, we investigated whether large-scale language models (LLMs) can help support decision-making processes based on their medical knowledge. Although similar reports already exist [1, 2], by including investigations in languages other than English, we aim to discuss the characteristics of LLMs in greater depth.

## Materials and methods

We used the brain MRI protocols that are employed at our hospital as a standard. These protocols include 20 disease names or examination purposes, including screening. However, this time, we did not include brain tumor protocols in this experiment because our hospital utilizes advanced and specialized imaging methods for brain tumors, such as navigation protocols, which are not suitable for evaluation as general sequences. We used the latest model of Generative Pretrained Transformer 4 (GPT-4) (OpenAI Inc., San Francisco, CA, USA), namely GPT-4o (gpt-4o-2024-05-13) [3]. We also used an in-house application to make sure that the data sent would not be used for GPT’s training. The temperature parameter, which controls the diversity of the results, was set to 0.0 to obtain a deterministic output. Messages sent to GPT consist of system prompts and user prompts, and the main part of instructions to GPT were described in the system prompt as shown below. Only the disease name was sent as the user prompt. We conducted this experiment in both Japanese and English and obtained 40 protocols from GPT. Our brain MRI protocol includes an axial T1-weighted image (T1WI Ax), axial T2-weighted image (T2WI Ax), axial T2-weighted fluid-attenuated inversion recovery (FLAIR Ax), axial diffusion-weighted image (DWI Ax), and time-of-flight magnetic resonance angiography (TOF MRA) as the standard set, with the option to add one additional sequence if necessary. We also asked GPT-4 to suggest only one additional sequence.

### System prompt for GPT-4 (Written in both English and Japanese)

> “*This GPT will assist in determining the MRI sequences for brain imaging. The basic imaging methods are T1WI Ax, T2WI Ax, FLAIR Ax, DWI Ax, and TOF MRA, and these should be performed in all cases. For the given disease name, if there is an additional sequence that needs to be imaged, please suggest only one. Since we do not want to increase the number of unnecessary tests, if the necessary sequence is included in the above, it is okay not to add any additional sequences. Do not include contrast-enhanced examinations. Do not include examinations of areas other than the brain*.

### In making the suggestion, please follow these steps

1. *Describe the MRI features of the disease*.
2. *Based on those features, select one additional imaging method to be added*.”

The protocols suggested by GPT were evaluated by two radiologists with over 20 years of relevant experience working together. The responses from GPT-4 were scored as follows: 0: Inappropriate, 1: Acceptable but does not match protocol used in our institute, 2: Matches our protocol.

## Result

GPT-4 suggested additional sequences for 17 of the 20 disease names in English and 13 sequences in Japanese. The scores for the sequences proposed by GPT were 27/40 points in English and 28/40 points in Japanese. GPT-4 provided inappropriate suggestions for Moyamoya disease and neuromyelitis optica in both languages and for cerebral infarction in Japanese. For the other protocols, the suggested sequences were either appropriate or better. The list of suggested sequences is shown in the table 1. Because the responses from GPT-4 were long sentences that included features of imaging findings, the full text is provided as supplementary information.

**Table 1.** Comparison of our sequences and GPT’s suggestions. Protocol names or target disease names, additional sequences used at our facility, and GPT’s suggestions in English and Japanese are presented. Some names are listed only as abbreviations. Because the responses from GPT-4 were long sentences that included the features of imaging findings, the full text is provided as supplementary information.

| Protocol and disease | Our sequence | GPT’s suggestion in English | GPT’s suggestion in Japanese |
|---|---|---|---|
| Cerebral aneurysm | None | None | None |
| Hemorrhage | T2*WI Ax | SWI | T2*WI Ax |
| Cavernous hemangioma | T2*WI Ax | GRE or SWI | SWI Ax |
| Arteriovenous malformation (AVM) | None | SWI | None |
| Moyamoya disease | Heavy T2 | SWI | None |
| Cerebral infarction | None | SWI | ADC map |
| Headache | None | None | None |
| Screening | None | None | None |
| Multiple sclerosis | FLAIR Cor | T2WI Sag | T2WI Sag |
| Neuromyelitis optica | FLAIR Cor | T2WI Cor | T2WI Sag |
| Alzheimer’s disease | FLAIR Cor | T1WI Cor | T1WI Cor |
| Epilepsy | FLAIR Cor | T2WI Cor or FLAIR Cor | T2WI Cor |
| Hydrocephalus | FLAIR Cor | Cine Phase-Contrast -MRI | None |
| Parkinson’s disease | T1WI Sag | SWI | SWI |
| Infective endocarditis, Sepsis | T2*WI Ax | SWI | T2*WI |
| Trigeminal neuralgia | DRIVE | 3D CISS or FIESTA | CISS or FIESTA |
| Hemifacial spasm | DRIVE | 3D CISS/FIESTA | CISS or FIESTA |
| Glossopharyngeal neuralgia | DRIVE | CISS or FIESTA | 3D T2WI |
| Spinocerebellar degeneration | T1WI Sag | T1WI Sag | T1WI Sag |
| Cerebellar ataxia, Cerebellar atrophy | T1WI Sag | T1WI Sag | T1WI Sag |

Here is a sample of GPT-4’s reply suggesting a sequence for cerebral aneurysm:

> **“1. MRI Imaging Features of Cerebral Aneurysm:**
>
> *- Cerebral aneurysms are often identified as saccular outpouchings of blood vessels in the brain. They may appear as flow voids on standard MRI sequences due to the rapid flow of blood within the aneurysm. The surrounding brain tissue may appear normal unless there has been a rupture, in which case there may be evidence of subarachnoid hemorrhage*.
>
> **2. Additional Imaging Method:**
>
> *- 3D TOF MRA (Time-of-Flight Magnetic Resonance Angiography): This sequence is particularly useful for visualizing blood vessels and detecting aneurysms. It provides high-resolution images of the cerebral vasculature and can help in assessing the size, shape, and location of the aneurysm. Since TOF MRA is already included in the basic imaging methods, no additional sequence is necessary*.”

## Discussion

Attempts to determine MRI sequences using natural language processing have been made with training using large datasets [4-6]; however, with the advent of pretrained models like GPT, we can now achieve this without the need for separate training [1, 2]. Nazario-Johnson et al. examined extensive neuroradiology protocols based on the American College of Radiology Appropriateness Criteria (ACR AC) [1]. However, the ACR AC is not applicable in our region. GPT-4 generally proposed appropriate additional sequences and explained the imaging findings supporting these suggestions in each language. In a preliminary experiment conducted before this study, the protocols generated freely by GPT-4 were more diverse, and the results were unstable. The most effective approach was to force GPT-4 to recall the disease’s imaging findings. This method significantly constrained the subsequent sequence suggestions, resulting in more stable outcomes. This approach is an example of prompt engineering known as the chain of thought [7].

Because of the limited time available for MR examinations at our institution, we are only allowed to add one sequence to the standard protocol. This limitation further constrained GPT-4’s output, which may have led to more favorable outcomes. According to GPT, TOF MRA is important for several protocols; however, it did not include this suggestion as it is already part of our basic protocol. This demonstrates that LLMs can handle complex decisions written in natural language.

GPT-4 made several inappropriate suggestions. For neuromyelitis optica, GPT-4 suggested coronal T2WI to observe optic nerve lesions. However, to accurately evaluate the optic nerves, it is preferable to suppress the signal from the orbital fat. Therefore, when imaging the orbits, the sequence should be T2WI with fat suppression or short inversion time inversion recovery (STIR)[8].

Moyamoya disease causes narrowing of the lumens of the internal carotid and middle cerebral arteries, and it is distinguished from arteriosclerosis by arterial shrinkage [9-11]. In Japan, where Moyamoya disease is more common, the diagnostic criteria for MRI include arterial vessel shrinkage on heavy T2WI [12]. GPT did not seem to have learned this diagnostic criterion even in Japanese.

GPT-4 suggested an ADC map for cerebral infarction in Japanese. The ADC map is generated alongside DWI, so it requires no additional sequence.

GPT-4’s suggested “3D CISS or 3D FIESTA” and “3D T2WI” for three protocols. The sequence actually used at our institute is spin-echo-based DRIVE (Philips Healthcare, Best, The Netherlands), which has the advantage of having fewer susceptibility artifacts compared with gradient-echo-based sequences such as CISS (Siemens Healthcare, Erlangen, Germany) or FIESTA (Philips) [13]. Since all these sequences are heavily T2-weighted 3D MR cisternography, they were scored as correct suggestions.

The sequences used in our institution were used as the gold standard for scoring. The training data of LLMs vary by language, causing their outputs to be language-dependent [14]. This approach offers utility tailored to specific regions and domains but introduces biases that hinder uniformity. Our results showed that the suggestions in English differed from those in Japanese for seven protocols. GPT-4 suggested SWI for six protocols in English, and T2*WI for two protocols along with SWI for two protocols in Japanese. Our actual protocol did not include SWI but included T2*WI for three protocols, which ameliorated the score in Japanese. SWI is superior to T2*WI in sensitivity to hemorrhage [15-17]. However, in our country, T2*WI is often preferred over SWI because the depiction of normal vessels in SWI can sometimes obscure hemorrhage site detection. In practice, neuroradiologists often make more complex decisions by reviewing images obtained during examinations and by referring to clinical information. The correct protocol may vary by region, facility, individual physician or radiologist, and patient, making it challenging for an LLM to provide a universally correct answer. Although this study focused solely on GPT, it is anticipated that other LLMs using the same Transformer algorithm will exhibit similar behaviors.

This experiment has some limitations. It only targeted head protocols, excluding brain tumors, and was limited to diseases that are commonly encountered at our hospital. The number of protocols tested was small, limiting the statistical validation. Restricting the model to suggest only one additional sequence and excluding the use of contrast agents are also limitations. We do not place significant emphasis on these issues because the purpose of this study is to explore the intrinsic behavior of LLMs and their potential role within the radiologist’s workflow.

Currently, LLM use for clinical decision-making is restricted by both regulatory and ethical guidelines. Please note that this report is based on a preliminary experiment and is not intended for clinical application. Further research on the radiological application of LLMs is needed.

## Conclusion

GPT-4 can suggest almost proper MRI sequences for each disease in addition to the standard brain MRI protocol. GPT-4’s output is language-dependent and suggests brain MRI protocols tailored to specific regions and domains.

## Statements and Declarations

## Acknowledgments

None.

## Ethical considerations

This article does not contain any studies with human or animal participants, therefore ethics approval was not required.

## Data availability

For complete results, including those in Japanese, please contact the corresponding author.

